# Comparative analyses of Alzheimer’s disease blood biomarkers and cognitive domains

**DOI:** 10.64898/2026.05.03.26352316

**Authors:** Deirdre M. O’Shea, James E. Galvin

## Abstract

**INTRODUCTION:** Whether Alzheimer’s disease (AD) blood biomarker-cognition associations differ across cognitive domains, analytic context, and biomarker modeling strategy in population-based cohorts is unclear.

**METHODS:** In 1,170 older adults from the Health and Retirement Study Harmonized Cognitive Assessment Protocol, we examined cross-sectional (2016) and prospective (2016-2022) associations of blood p-tau181, glial fibrillary acidic protein (GFAP), neurofilament light (NfL), and amyloid-β42/40 with memory, executive function, language, visuospatial ability, and global cognition using individual biomarker, principal components analysis-derived composite, and multibiomarker panel models.

**RESULTS:** Cross-sectionally, NfL and GFAP showed the broadest associations. Prospectively, p-tau181 was independently associated with memory and global cognition, whereas GFAP was associated with executive function, memory, and global cognition. P-tau181 also showed relative memory-versus-executive selectivity. The comparatively best-fitting modeling approach differed by cognitive domain and analytic context.

**DISCUSSION:** AD blood biomarker-cognition associations in community-dwelling older adults are domain-differentiated and context-dependent, supporting domain-specific outcomes and flexible biomarker modeling strategies.

## 1 Introduction

Blood-based biomarkers (BBMs) have emerged as scalable tools for detecting Alzheimer’s disease (AD) and related neurodegenerative processes^1^. Among them, the amyloid-β42/40 ratio (Aβ42/40) and phosphorylated tau-181 (p-tau181) index core AD proteinopathies, whereas neurofilament light chain (NfL) and glial fibrillary acidic protein (GFAP) index neuroaxonal injury and astrocytic activation, respectively.^2–6^ These markers have shown prognostic associations with subsequent cognitive decline, including in cognitively unimpaired older adults,^7–9^ but most prognostic work has relied on global cognitive composites as outcomes and clinic-based cohorts enriched for cognitive impairment^10–12^. However, whether these associations generalize to population-based settings remains uncertain, as mixed pathologies and comorbidity may attenuate biomarker-cognition relationships^13–15^. These settings may nonetheless be where BBMs have greatest public health value as scalable tools for probabilistic risk stratification and triage.

Global cognitive composites may have limited sensitivity to early biomarker-associated cognitive changes, which in AD often appear first in episodic memory before extending to other domains.^16,17^ In population-based samples, where biomarker–cognition associations may be more heterogeneous than in clinic-based cohorts, reliance on global cognitive composites may obscure domain-differentiated associations among biomarkers indexing distinct biological processes.^14,18,19^ Domain-specific outcomes may therefore be more sensitive to biomarker-cognition associations that precede detectable global impairment and may clarify the clinical interpretation of biomarker results, for example by distinguishing an AD-memory-predominant profile from a broader pattern of multidomain vulnerability. Several population-based or community-based studies have examined domain-specific BBM-cognition associations. In the Monongahela-Youghiogheny Healthy Aging Team study (MYHAT), p-tau181 showed the strongest cross-sectional associations with memory, whereas NfL was more strongly associated with attention and executive performance; prospectively, Aβ42/40 was most strongly associated with memory change and GFAP with language change.^18^ In the HABS-HD, higher plasma p-tau181 and NfL were associated cross-sectionally with multiple cognitive domains, including memory, executive function, processing speed, and language, in a racially and ethnically diverse community-based sample.^20^ While in the UK Biobank study, increases in GFAP longitidinally were associated with decline in processing speed and visual attention ^21^. Other studies have similarly shown that plasma biomarkers predict incident dementia over long follow-up periods, ^22^ yet most prognostic analyses continue to rely on global cognitive composites rather than domain-specific outcomes.

Although these studies support heterogeneity in biomarker-cognition associations, important gaps remain. First, although prior studies have described domain-level patterns, few have formally tested whether a given biomarker’s association with one cognitive domain differs statistically from its association with another. Without such tests, apparent domain selectivity may reflect sampling variability; more generally, a significant association in one domain and a nonsignificant association in another does not establish that the two associations differ significantly.^23^ Establishing whether a biomarker is linked preferentially to memory versus broader cognitive functioning would strengthen interpretation of biomarker profiles, guide selection of cognitive outcomes, and inform targeting of monitoring strategies and prevention trials. Second, individual biomarker models, composites, and simultaneous panels have each been applied,^7,12,24,25^ but they have rarely been compared systematically against the same domain-specific outcomes within the same cohort. Whether the optimal modeling approach differs by cognitive domain therefore remains unclear.

Third, whether the optimal biomarker combination differs depending on whether the goal is to characterize concurrent cognitive status or predict baseline-adjusted follow-up cognition has not been tested. If different markers predominate at different stages of disease progression ^26^, the most parsimonious biomarker configuration may differ across cross-sectional and prospective applications, with implications for screening, prognostic enrichment, and prevention trial design.

Together, these gaps align with recent calls for context-aware interpretation frameworks for plasma biomarkers that account for analytic context, population heterogeneity, and intended use.^27^ The current study therefore examined whether BBMs show domain-differentiated cognitive associations, whether these associations differ across cross-sectional and prospective analyses, and whether relative domain selectivity and model performance vary across biomarker approaches in a population-based sample of older U.S. adults.

## 2. Methods

### 2.1 Study Population and Design

Data were drawn from the 2016 and 2022 waves of the Harmonized Cognitive Assessment Protocol (HCAP),^28^ a sub-study of the Health and Retirement Study (HRS).^29^ The HRS is an ongoing nationally representative, biennial longitudinal panel study of US adults aged 51 and older that employs a stratified, multistage probability sampling design with oversampling of Black, Hispanic, and high-wealth households. The HCAP was designed to facilitate international harmonization of cognitive assessment and to characterize the prevalence and determinants of cognitive impairment in the US older adult population.^28^ Of the 1,657 participants with cognitive data at both the 2016 and 2022 HCAP waves, 1,287 had complete blood biomarker data. After excluding participants with missing covariate data, the final analytic sample comprised 1,170 individuals.

### 2.2 Measures

#### 2.2.1 Cognitive Assessment

Cognitive test administration was performed in participants’ homes, which included an approximately 1-hour respondent interview and a 20-minute informant interview administered by trained interviewers.^28^ The test battery incorporated items from brief mental-status instruments, including the Mini-Mental State Examination (MMSE),^30^ Telephone Interview for Cognitive Status (TICS),^31^ and Community Screening Instrument for Dementia (CSI-D), ^32^ together with neuropsychological tasks assessing memory, language, executive functioning, and visuospatial construction.

Domain composites were created a priori by averaging available z-scored standardized indicators within each domain for each time point. Episodic memory was indexed using delayed and recognition measures drawn from the CERAD word-list and constructional praxis tasks, ^33^ MMSE three-word delayed recall,^30^ delayed story recall from the East Boston Memory Test, ^34^ and delayed story memory from the Wechsler Memory Scale Logical Memory subtest. ^35^ Executive function was indexed using Trail Making Test Parts A and B,^36^ backward counting from the Brief Test of Adult Cognition by Telephone,^37^ Raven’s Standard Progressive Matrices, ^38^ and the HRS Number Series task. ^39^ The Trail Making Test Part A and Part B time scores were log-transformed and reverse-scored so that higher values indicated better performance. Language was indexed by animal naming, adapted from the Woodcock–Johnson III^40^ and confrontation-naming items drawn from the TICS, MMSE, and CSI-D.^30–32^ Visuospatial ability was assessed with constructional praxis copy from the CERAD battery, ^33^ the MMSE copy polygons was excluded due to psychometric redundancy with CERAD in the primary HRS-HCAP factor analytic work.^41^ Domain scores were computed from available indicators without imputation.

A global cognition composite was derived at baseline (2016) by extracting the first unrotated principal component from all item-level z-scores, with median imputation applied to address limited item-level missingness. To maintain comparability across waves, the 2022 global cognition score was derived by applying the 2016 PCA loadings to the standardized 2022 item scores and rescaling the resulting composite to the 2016 metric (mean = 50, SD = 10). Baseline cognitive status (cognitively normal [CN], mild cognitive impairment [MCI], dementia) was assigned using the published HRS-HCAP adjudication algorithm for the 2016 wave.^42^

#### 2.2.2 Blood-Based Biomarkers

Blood samples were collected as part of the 2016 HRS Venous Blood Study,^43^ with venous draws performed in participants’ homes by certified phlebotomists. Samples were shipped overnight in temperature-controlled packaging to the Advanced Research and Diagnostics Laboratory at the University of Minnesota. Prior validation studies have confirmed the stability of this protocol for quantifying neuropathological biomarkers, with 92% of samples received within 24 hours and 99% within 48 hours.^44^

As described in the HRS documentation for this study,^45^ blood plasma biomarkers included Aβ42/40 ratio, GFAP, and NfL, were measured in EDTA plasma using a Simoa HD-X analyzer with the Neurology 4-plex assay E (N4PE; Quanterix).^46^ Serum p-tau181 was measured using the Simoa pTau-181 V2 assay (Quanterix). All assays were conducted at the University of Minnesota Advanced Research and Diagnostic Laboratory. Biomarker values were natural log-transformed prior to z-standardization. For the Aβ42/40 ratio that lower z-scores reflect greater amyloid burden, consistent with the conventional interpretation of the ratio whereby a lower Aβ42/40 value indicates greater amyloid plaque accumulation.

#### 2.2.3 APOE Genotyping

Described in the HRS study protocol,^47^ APOE genotypes were directly assayed on salivary DNA using TaqMan allelic discrimination assays targeting rs429358 and rs7412 on a 7900HT Sequence Detection System; when direct APOE genotypes were unavailable, APOE status was derived from imputed genome-wide genotype data generated from Illumina HumanOmni2.5 arrays using the 1000 Genomes Project Phase 3 worldwide reference panel. Genome-wide genotyping was performed at the Center for Inherited Disease Research at Johns Hopkins University. APOE status was dichotomized as ε4 carrier versus non-carrier.

### 2.3 Statistical Analyses

All analyses were conducted in R (version 4.5.2; R Core Team, 2025). Descriptive characteristics were summarized using means and standard deviations for continuous variables and frequencies and percentages for categorical variables. Bivariate associations among biomarkers were evaluated using Spearman rank correlations, and biomarker distributions were examined by race/ethnicity using group means and Kruskal-Wallis tests. Attrition analyses compared the baseline-eligible sample to the final analytic sample on demographics, APOE ε4 status, and biomarker levels.

#### 2.3.1 Primary Biomarker Models

Three primary modeling strategies were evaluated: (1) individual biomarker models, in which each biomarker was entered separately; (2) a PCA-derived biomarker composite, in which the first unrotated principal component from the four standardized biomarkers was entered as a single predictor; and (3) a multivariable panel, in which all four biomarkers were entered simultaneously.

#### 2.3.2 Cross-Sectional and Prospective Association Models

Cross-sectional associations between 2016 biomarkers and concurrent 2016 cognitive performance were examined using multiple linear regression. Prospective associations between 2016 biomarkers and 2022 cognitive performance were examined using analysis of covariance (ANCOVA), with the corresponding 2016 domain score included as a covariate, thereby indexing 2022 performance conditional on baseline. All models adjusted for age, sex, years of education, race/ethnicity (non-Hispanic White [reference], non-Hispanic Black, Hispanic, other non-Hispanic), and APOE ε4 carrier status. All continuous predictors and outcomes were standardized to z-scores (βs).

#### 2.3.3 Model Comparison and Cross-Sectional–Prospective Coefficient Differences

Comparative model fit was evaluated using the Akaike Information Criterion (AIC) and Bayesian Information Criterion (BIC), with lower values indicating better fit ^48^. Relative to AIC, BIC imposes a stronger penalty for model complexity and was therefore interpreted as a more stringent index of parsimony. Nested-model F-tests compared each biomarker model with a restricted covariate-only null model (1 df for individual and PCA models; 4 df for the full panel). Incremental variance explained (ΔR²) was quantified relative to the null model. To compare biomarker-cognition associations between the cross-sectional and prospective analyses, standardized coefficients from the multivariable models were compared for each biomarker-domain combination using z-tests, defined as the difference in coefficients divided by the square root of the summed squared standard errors. P-values were FDR-corrected across comparisons.

#### 2.3.4 Relative Memory-Versus-Executive Selectivity and Reduced Composite Analyses

Analyses were conducted separately for each biomarker. Relative memory-versus-executive selectivity was evaluated using stacked mixed-effects models including biomarker tertile, domain, and a tertile × domain interaction, adjusted for age, sex, education, race/ethnicity, APOE ε4 status, and the corresponding 2016 domain score, with a random intercept for participant. Interaction p-values were FDR-corrected across biomarkers. Planned Memory − Executive contrasts of the High-versus-Low tertile effect were examined as follow-up tests and similarly FDR-corrected across biomarkers.

Where primary multivariable results indicated overlapping biomarker contributions, follow-up exploratory model-selection analyses examined whether reduced biomarker composites could represent that information more parsimoniously than the full four-biomarker panel. Candidate composites were compared using ΔAIC and ΔBIC relative to the full panel, with negative values indicating improved fit with fewer parameters.

#### 2.3.5 Sensitivity Analyses

Primary models were re-estimated in two clinical subsamples: (a) participants without dementia (CN + MCI; n = 1,137) and (b) cognitively normal participants only (CN; n = 931). To examine whether prospective biomarker-cognition associations varied by race/ethnicity or APOE ε4 status, given reported differences in plasma biomarker levels and associations across these characteristics ^11,20^, interaction models were estimated by adding one biomarker × moderator term at a time to the prospective multivariable model. For race/ethnicity moderation, non-Hispanic White participants served as the reference group; participants identifying as Other, non-Hispanic (n = 22) were excluded from moderation analyses due to insufficient cell sizes but retained as a covariate category in all primary models. Omnibus race/ethnicity interaction F-tests (2 df) and APOE ε4 interaction F-tests (1 df) were FDR-corrected across all biomarker-domain combinations. Stratified coefficients by APOE ε4 carrier status were extracted to characterize the direction of significant interactions. Moderation analyses were conducted in the full analytic sample.

#### 2.3.6 Multiple Comparison Correction

P-values for individual regression coefficients were adjusted using the Benjamini-Hochberg procedure within each cognitive domain × model type combination where q < .05 indicated statistical significance ^49^. F-test p-values were similarly adjusted across cognitive domains. FDR correction for tertile and domain selectivity analyses was applied across all biomarker-domain combinations; for moderation analyses, correction was applied across all biomarker-domain interaction tests within each moderator.

## 3. Results

### 3.1 Sample Characteristics

The analytic sample comprised 1,170 participants (931 CN, 206 MCI, 33 dementia; **Table 1**). Participants were 73.4 ± 6.0 years old with 13.2 ± 3.0 years of education; 58.9% were female. The sample was 72.6% non-Hispanic White, 14.0% non-Hispanic Black, 11.5% Hispanic, and 1.9% other non-Hispanic. APOE ε4 carriers constituted 23.6% of the sample. Attrition analyses revealed no significant differences between the baseline-eligible sample (N = 1,287) and the retained analytic sample on demographics, APOE ε4 status, or biomarker levels (all p > .40; Supplementary **Table S1**).

**Table 1.**
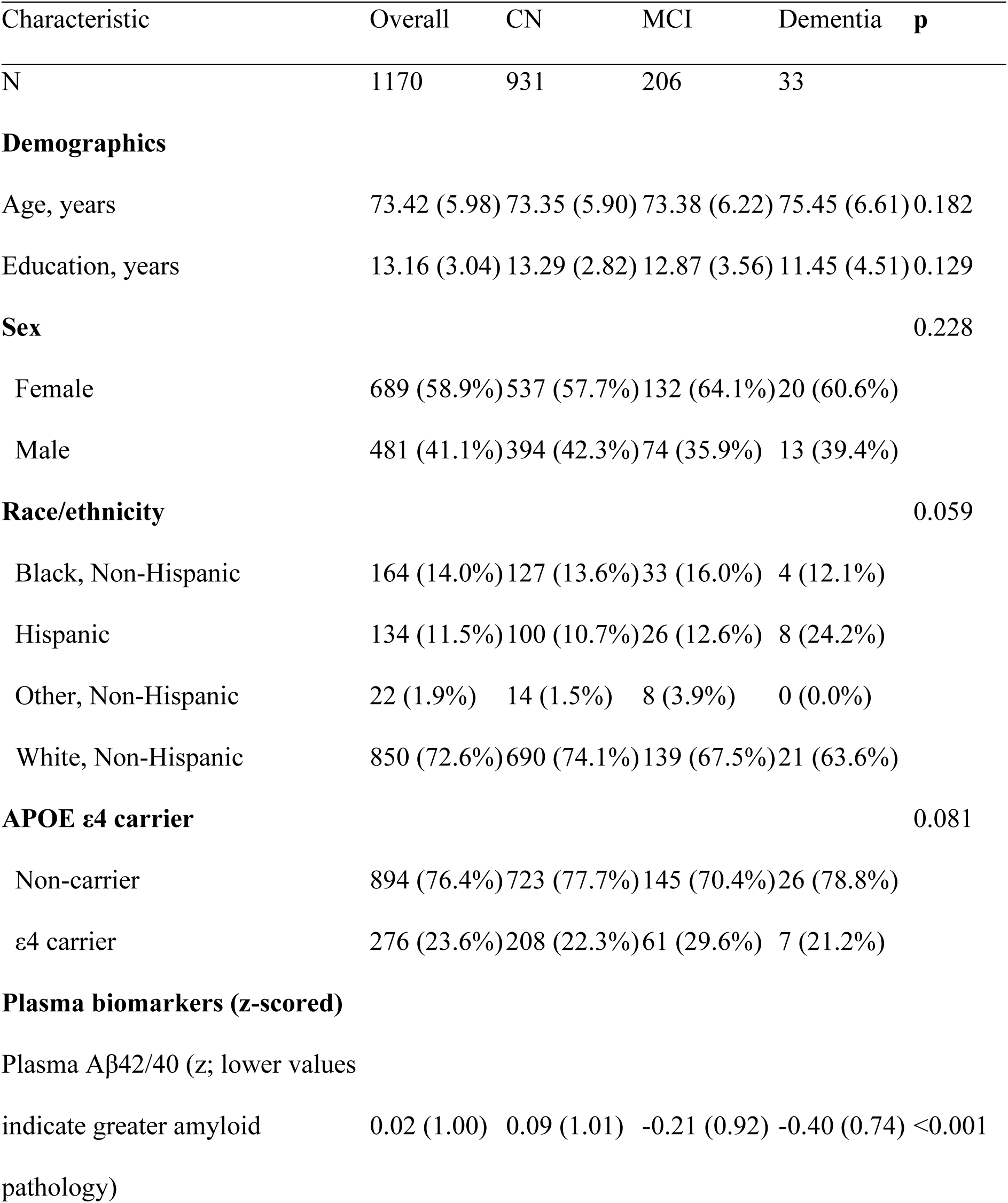
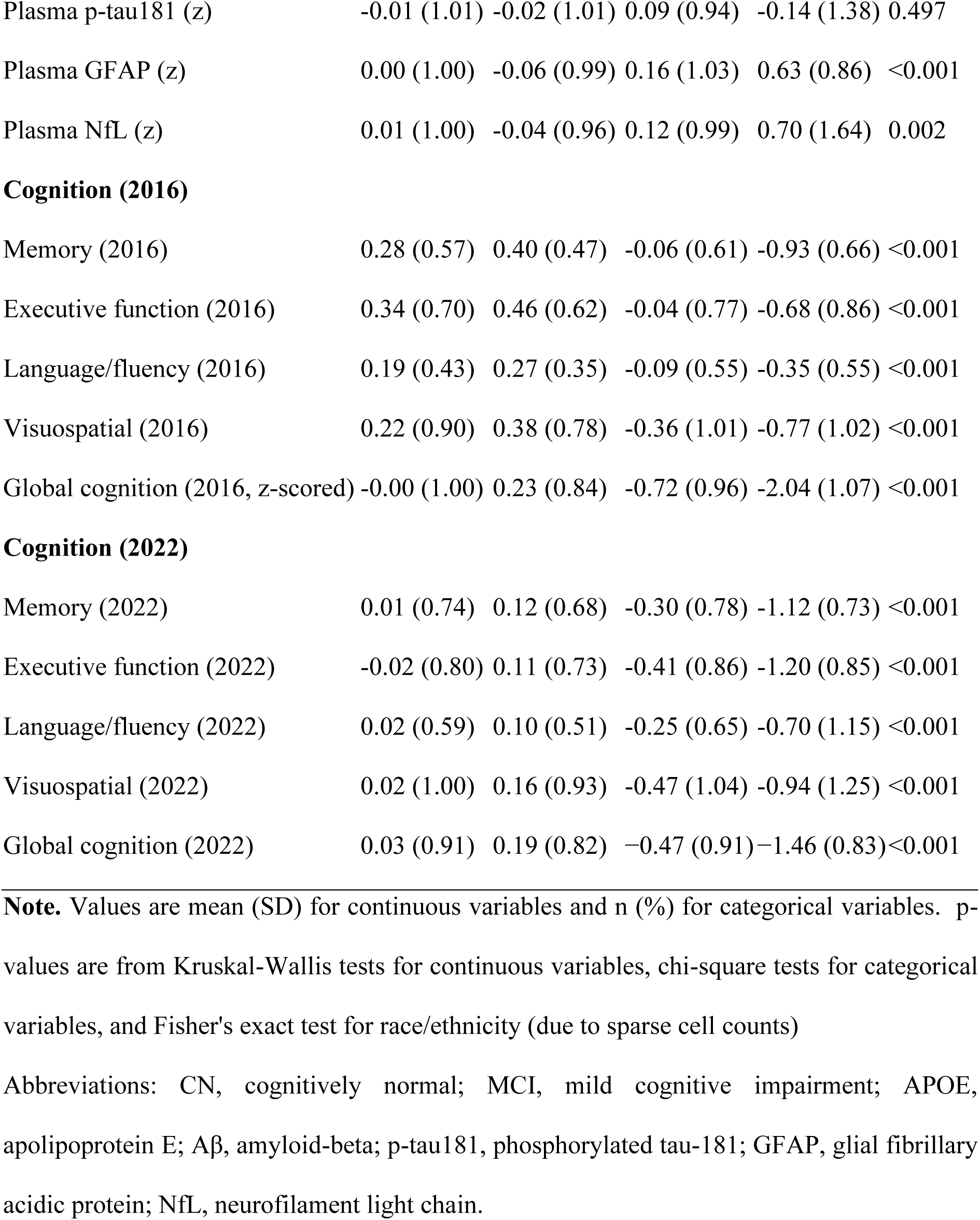
Characteristics of analytic sample (N=1170) by baseline (HCAP 2016) cognitive status.

| Characteristic | Overall | CN | MCI | Dementia | p |
| --- | --- | --- | --- | --- | --- |
| N | 1170 | 931 | 206 | 33 |  |
| **Demographics** |  |  |  |  |  |
| Age, years | 73.42 (5.98) | 73.35 (5.90) | 73.38 (6.22) | 75.45 (6.61) | 0.182 |
| Education, years | 13.16 (3.04) | 13.29 (2.82) | 12.87 (3.56) | 11.45 (4.51) | 0.129 |
| **Sex** |  |  |  |  | 0.228 |
| Female | 689 (58.9%) | 537 (57.7%) | 132 (64.1%) | 20 (60.6%) |  |
| Male | 481 (41.1%) | 394 (42.3%) | 74 (35.9%) | 13 (39.4%) |  |
| **Race/ethnicity** |  |  |  |  | 0.059 |
| Black, Non-Hispanic | 164 (14.0%) | 127 (13.6%) | 33 (16.0%) | 4 (12.1%) |  |
| Hispanic | 134 (11.5%) | 100 (10.7%) | 26 (12.6%) | 8 (24.2%) |  |
| Other, Non-Hispanic | 22 (1.9%) | 14 (1.5%) | 8 (3.9%) | 0 (0.0%) |  |
| White, Non-Hispanic | 850 (72.6%) | 690 (74.1%) | 139 (67.5%) | 21 (63.6%) |  |
| **APOE ε4 carrier** |  |  |  |  | 0.081 |
| Non-carrier | 894 (76.4%) | 723 (77.7%) | 145 (70.4%) | 26 (78.8%) |  |
| ε4 carrier | 276 (23.6%) | 208 (22.3%) | 61 (29.6%) | 7 (21.2%) | |
| **Plasma biomarkers (z-scored)** |  |  |  |  |  |
| Plasma Aβ42/40 (z; lower values indicate greater amyloid pathology) | 0.02 (1.00) | 0.09 (1.01) | -0.21 (0.92) | -0.40 (0.74) | <0.001 |
| Plasma p-tau181 (z) | -0.01 (1.01) | -0.02 (1.01) | 0.09 (0.94) | -0.14 (1.38) | 0.497 |
| Plasma GFAP (z) | 0.00 (1.00) | -0.06 (0.99) | 0.16 (1.03) | 0.63 (0.86) | <0.001 |
| Plasma NfL (z) | 0.01 (1.00) | -0.04 (0.96) | 0.12 (0.99) | 0.70 (1.64) | 0.002 |
| **Cognition (2016)** |  |  |  |  |  |
| Memory (2016) | 0.28 (0.57) | 0.40 (0.47) | -0.06 (0.61) | -0.93 (0.66) | <0.001 |
| Executive function (2016) | 0.34 (0.70) | 0.46 (0.62) | -0.04 (0.77) | -0.68 (0.86) | <0.001 |
| Language/fluency (2016) | 0.19 (0.43) | 0.27 (0.35) | -0.09 (0.55) | -0.35 (0.55) | <0.001 |
| Visuospatial (2016) | 0.22 (0.90) | 0.38 (0.78) | -0.36 (1.01) | -0.77 (1.02) | <0.001 |
| Global cognition (2016, z-scored) | -0.00 (1.00) | 0.23 (0.84) | -0.72 (0.96) | -2.04 (1.07) | <0.001 |
| **Cognition (2022)** |  |  |  |  |  |
| Memory (2022) | 0.01 (0.74) | 0.12 (0.68) | -0.30 (0.78) | -1.12 (0.73) | <0.001 |
| Executive function (2022) | -0.02 (0.80) | 0.11 (0.73) | -0.41 (0.86) | -1.20 (0.85) | <0.001 |
| Language/fluency (2022) | 0.02 (0.59) | 0.10 (0.51) | -0.25 (0.65) | -0.70 (1.15) | <0.001 |
| Visuospatial (2022) | 0.02 (1.00) | 0.16 (0.93) | -0.47 (1.04) | -0.94 (1.25) | <0.001 |
| Global cognition (2022) | 0.03 (0.91) | 0.19 (0.82) | -0.47 (0.91) | -1.46 (0.83) | <0.001 |

**Note.** Values are mean (SD) for continuous variables and n (%) for categorical variables. p-values are from Kruskal-Wallis tests for continuous variables, chi-square tests for categorical variables, and Fisher's exact test for race/ethnicity (due to sparse cell counts)
Abbreviations: CN, cognitively normal; MCI, mild cognitive impairment; APOE, apolipoprotein E; Aβ , amyloid-beta; p-tau181, phosphorylated tau-181; GFAP, glial fibrillary acidic protein; NfL, neurofilament light chain.

Expected gradients across diagnostic groups were observed for all cognitive domains at both time points (all p < .001). Among biomarkers, lower Aβ42/40 and higher GFAP and NfL were observed in the dementia group (p ≤ .002); p-tau181 did not differ significantly across groups (p = .497). Biomarker distributions differed significantly by race/ethnicity for p-tau181, NfL, and Aβ42/40 (all p < .001) but not GFAP (p = .149; Supplementary **Table S2**). Non-Hispanic Black participants showed lower NfL and Aβ42/40 relative to non-Hispanic White participants; Hispanic participants showed the lowest p-tau181. APOE ε4 carrier prevalence was highest among non-Hispanic Black participants (34.8%) compared with non-Hispanic White (21.9%) and Hispanic (19.4%) participants. Biomarker correlations are presented in Supplementary **Table S3.**

### 3.2 Biomarker PCA

The first unrotated principal component loaded strongly on NfL (0.86), GFAP (0.80), and p-tau181 (0.62), with a near-zero loading for Aβ42/40 (−0.06), indicating that the PCA biomarker composite primarily captured shared neurodegeneration and tau variance with negligible amyloid contribution.

### 3.3 Cross-sectional Associations

Full Cross-sectional results are reported in Supplementary **Table S4**. In brief, NfL and GFAP showed the broadest associations with cross-sectional cognitive performance in individual biomarker models, with significant negative associations across memory, executive function, and global cognition (all q < .001). Aβ42/40 was positively associated with executive function, global cognition, and language (q ≤ .013), reflecting better performance with lower amyloid burden. P-tau181 associations were modest and largely attenuated after FDR correction. In multivariable models, NfL and GFAP effects remained robust while p-tau181 was no longer independently significant.

### 3.4 Prospective Associations

#### 3.4.1 Individual Biomarker Models

Full prospective results are summarized in **Table 2**. In contrast to the cross-sectional pattern, p-tau181 showed the strongest prospective associations with memory (β = −0.088, 95% CI [−0.119, −0.057], q < .001) and global cognition (β = −0.078, q < .001), with a modest association with executive function (β = −0.028, q = .033). GFAP was associated with executive function (β = −0.060, q < .001), memory (β = −0.063, q < .001), and global cognition (β = −0.079, q < .001). NfL showed associations with executive function (β = −0.049, q < .001), memory (β = −0.049, q = .006), and global cognition (β = −0.055, q = .003). Aβ42/40 showed no significant prospective associations (all q > .26).

**Table 2.**
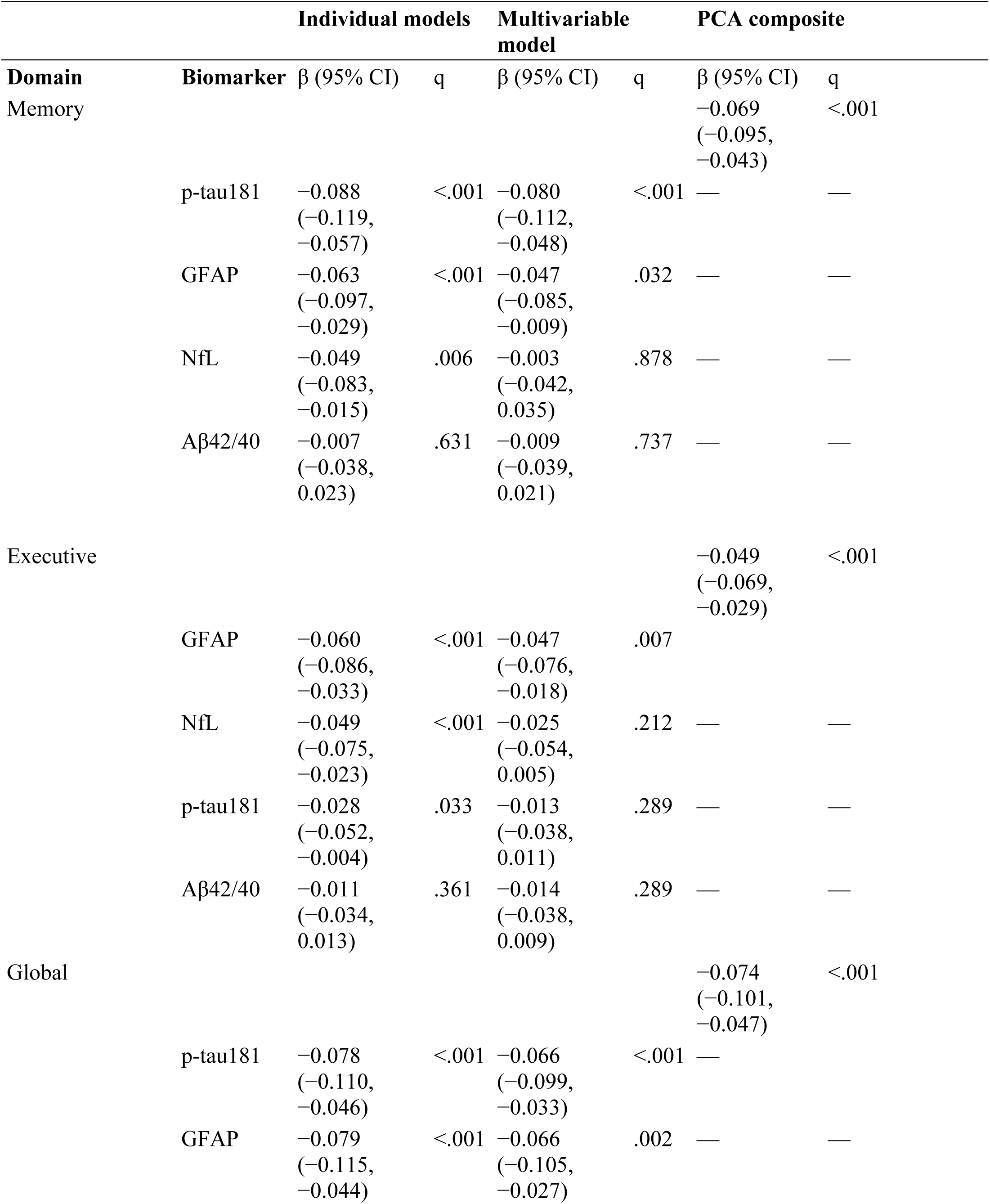
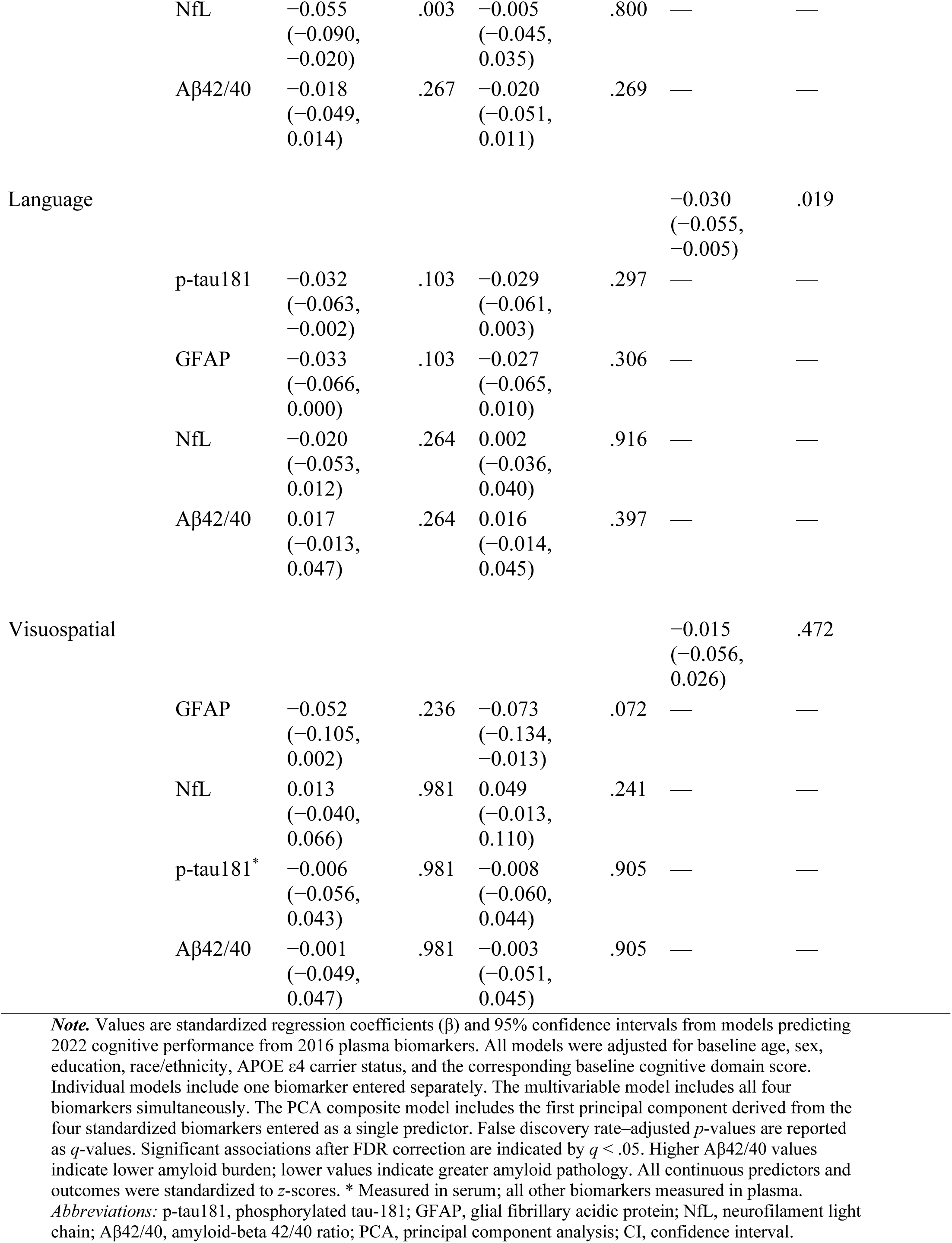
Prospective Associations of Baseline Plasma Biomarkers with Cognitive Performance 6 Years Later Across Cognitive Domains.

| Domain | Biomarker | Individual models |  | Multivariable model |  | PCA composite |  |
| --- | --- | --- | --- | --- | --- | --- | --- |
| | | $\beta$ (95% CI) | q | $\beta$ (95% CI) | q | $\beta$ (95% CI) | q |
| Memory |  |  |  |  |  | −0.069<br>(−0.095,<br>−0.043) | <.001 |
|  | p-tau181 | −0.088<br>(−0.119,<br>−0.057) | <.001 | −0.080<br>(−0.112,<br>−0.048) | <.001 | — | — |
|  | GFAP | −0.063<br>(−0.097,<br>−0.029) | <.001 | −0.047<br>(−0.085,<br>−0.009) | .032 | — | — |
|  | NfL | −0.049<br>(−0.083,<br>−0.015) | .006 | −0.003<br>(−0.042,<br>0.035) | .878 | — | — |
| | Aβ42/40 | −0.007<br>(−0.038,<br>0.023) | .631 | −0.009<br>(−0.039,<br>0.021) | .737 | — | — |
| Executive |  |  |  |  |  | −0.049<br>(−0.069,<br>−0.029) | <.001 |
|  | GFAP | −0.060<br>(−0.086,<br>−0.033) | <.001 | −0.047<br>(−0.076,<br>−0.018) | .007 |  |  |
|  | NfL | −0.049<br>(−0.075,<br>−0.023) | <.001 | −0.025<br>(−0.054,<br>0.005) | .212 | — | — |
|  | p-tau181 | −0.028<br>(−0.052,<br>−0.004) | .033 | −0.013<br>(−0.038,<br>0.011) | .289 | — | — |
| | Aβ42/40 | −0.011<br>(−0.034,<br>0.013) | .361 | −0.014<br>(−0.038,<br>0.009) | .289 | — | — |
| Global |  |  |  |  |  | −0.074<br>(−0.101,<br>−0.047) | <.001 |
|  | p-tau181 | −0.078<br>(−0.110,<br>−0.046) | <.001 | −0.066<br>(−0.099,<br>−0.033) | <.001 | — |  |
|  | GFAP | −0.079<br>(−0.115,<br>−0.044) | <.001 | −0.066<br>(−0.105,<br>−0.027) | .002 | — | — |
|  | NfL | −0.055<br>(−0.090,<br>−0.020) | .003 | −0.005<br>(−0.045,<br>0.035) | .800 | — | — |
|  | Aβ42/40 | −0.018<br>(−0.049,<br>0.014) | .267 | −0.020<br>(−0.051,<br>0.011) | .269 | — | — |
| Language |  |  |  |  |  | −0.030<br>(−0.055,<br>−0.005) | .019 |
|  | p-tau181 | −0.032<br>(−0.063,<br>−0.002) | .103 | −0.029<br>(−0.061,<br>0.003) | .297 | — | — |
|  | GFAP | −0.033<br>(−0.066,<br>0.000) | .103 | −0.027<br>(−0.065,<br>0.010) | .306 | — | — |
|  | NfL | −0.020<br>(−0.053,<br>0.012) | .264 | 0.002<br>(−0.036,<br>0.040) | .916 | — | — |
|  | Aβ42/40 | 0.017<br>(−0.013,<br>0.047) | .264 | 0.016<br>(−0.014,<br>0.045) | .397 | — | — |
| Visuospatial |  |  |  |  |  | −0.015<br>(−0.056,<br>0.026) | .472 |
|  | GFAP | −0.052<br>(−0.105,<br>0.002) | .236 | −0.073<br>(−0.134,<br>−0.013) | .072 | — | — |
|  | NfL | 0.013<br>(−0.040,<br>0.066) | .981 | 0.049<br>(−0.013,<br>0.110) | .241 | — | — |
|  | p-tau181* | −0.006<br>(−0.056,<br>0.043) | .981 | −0.008<br>(−0.060,<br>0.044) | .905 | — | — |
|  | Aβ42/40 | −0.001<br>(−0.049,<br>0.047) | .981 | −0.003<br>(−0.051,<br>0.045) | .905 | — | — |

**Note.** Values are standardized regression coefficients ( $\beta$ ) and 95% confidence intervals from models predicting 2022 cognitive performance from 2016 plasma biomarkers. All models were adjusted for baseline age, sex, education, race/ethnicity, APOE ε4 carrier status, and the corresponding baseline cognitive domain score. Individual models include one biomarker entered separately. The multivariable model includes all four biomarkers simultaneously. The PCA composite model includes the first principal component derived from the four standardized biomarkers entered as a single predictor. False discovery rate-adjusted $p$ -values are reported as $q$ -values. Significant associations after FDR correction are indicated by $q < .05$ . Higher Aβ42/40 values indicate lower amyloid burden; lower values indicate greater amyloid pathology. All continuous predictors and outcomes were standardized to $z$ -scores. \* Measured in serum; all other biomarkers measured in plasma. *Abbreviations:* p-tau181, phosphorylated tau-181; GFAP, glial fibrillary acidic protein; NfL, neurofilament light chain; Aβ42/40, amyloid-beta 42/40 ratio; PCA, principal component analysis; CI, confidence interval.

#### 3.4.2 Multivariable Panel Models

After mutual adjustment, p-tau181 retained independent associations with memory (β = −0.080, q < .001) and global cognition (β = −0.066, q < .001). GFAP retained independent associations with executive function (β = −0.047, q = .007), memory (β = −0.047, q = .032), and global cognition (β = −0.066, q = .002). NfL and Aβ42/40 were not independently significant for any domain (all q > .21). Results are shown in **Figure 1**.

**Figure 1.**
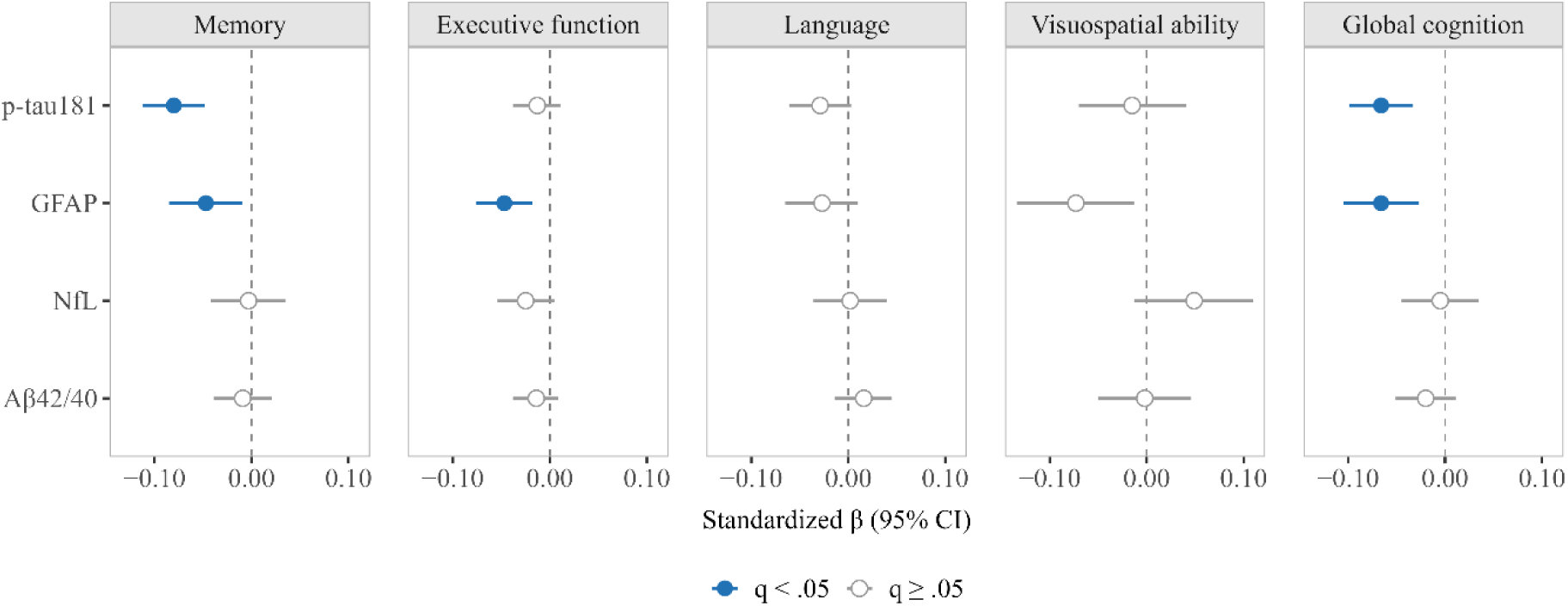
Multivariable associations of baseline plasma biomarkers with baseline-adjusted cognitive performance 6 years later across cognitive domains. *Note.* Points represent standardized regression coefficients (β) and error bars 95% confidence intervals from multivariable models including p-tau181, GFAP, NfL, and Aβ42/40 entered simultaneously. Models predict 2022 cognitive performance from 2016 plasma biomarkers, adjusted for baseline age, sex, education, race/ethnicity, APOE ε4 status, and the corresponding baseline cognitive domain score. All continuous predictors and outcomes were standardized to *z*-scores. Higher Aβ42/40 values indicate lower amyloid burden; lower values indicate greater amyloid pathology. False discovery rate–adjusted *p*-values are reported as *q*-values. Abbreviations: p-tau181, phosphorylated tau-181; GFAP, glial fibrillary acidic protein; NfL, neurofilament light chain; Aβ42/40, amyloid-beta 42/40 ratio; CI, confidence interval.

#### 3.4.3 PCA Composite

The PCA biomarker composite was significantly associated with memory (β = −0.069, q < .001), executive function (β = −0.049, q < .001), global cognition (β = −0.074, q < .001), and language (β = −0.030, q = .019), but not visuospatial performance (q = .472).

### 3.5 Primary Model Comparison

**Table 3** presents model fit indices for all three modeling strategies. Base models explained 28.0%–75.5% of variance in prospective cognitive performances (R² range), primarily reflecting the autoregressive effect of baseline cognition. Biomarkers explained a modest additional increment (ΔR² = .000–.016), with significant improvements for memory, executive function, global cognition, and language. The preferred modeling approach differed by domain: for memory, the multivariable panel was favored by AIC and individual p-tau181 by BIC; for executive function, the PCA biomarker composite was preferred by both criteria; for global cognition, the multivariable model by AIC and the PCA biomarker composite by BIC; biomarker contributions were marginal for language and minimal for visuospatial performance.

**Table 3.**
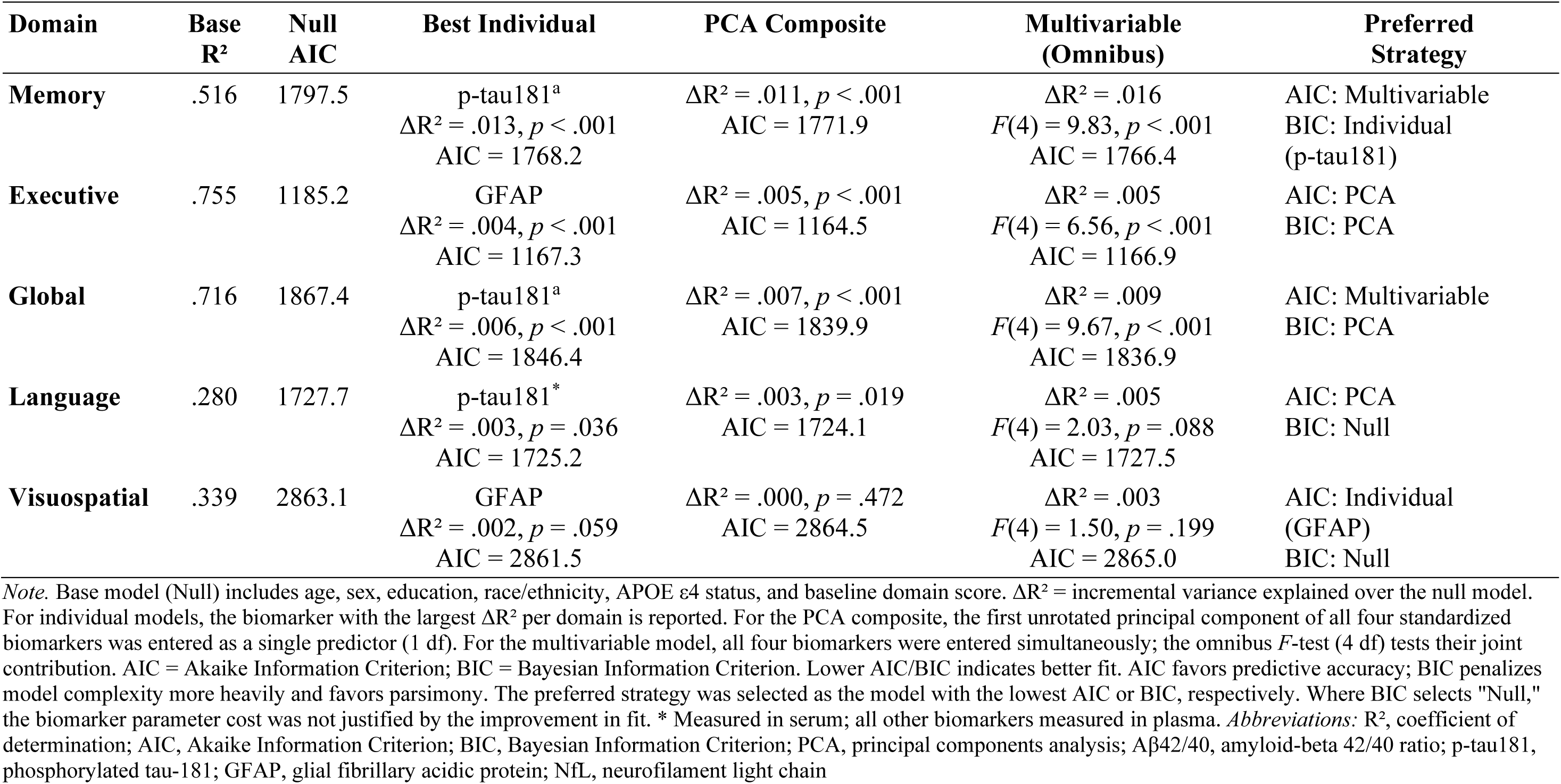
Model Comparison: Incremental Predictive Value of Three Biomarker Modeling Strategies for Prospective Cognitive Outcomes (Full Sample, N = 1,170)

| Domain | Base R<sup>2</sup> | Null AIC | Best Individual | PCA Composite | Multivariable (Omnibus) | Preferred Strategy |
| --- | --- | --- | --- | --- | --- | --- |
| **Memory** | .516 | 1797.5 | p-tau181<sup>a</sup><br>$\Delta R^2 = .013, p < .001$<br>AIC = 1768.2 | $\Delta R^2 = .011, p < .001$<br>AIC = 1771.9 | $\Delta R^2 = .016$<br>$F(4) = 9.83, p < .001$<br>AIC = 1766.4 | AIC: Multivariable<br>BIC: Individual (p-tau181) |
| **Executive** | .755 | 1185.2 | GFAP<br>$\Delta R^2 = .004, p < .001$<br>AIC = 1167.3 | $\Delta R^2 = .005, p < .001$<br>AIC = 1164.5 | $\Delta R^2 = .005$<br>$F(4) = 6.56, p < .001$<br>AIC = 1166.9 | AIC: PCA<br>BIC: PCA |
| **Global** | .716 | 1867.4 | p-tau181<sup>a</sup><br>$\Delta R^2 = .006, p < .001$<br>AIC = 1846.4 | $\Delta R^2 = .007, p < .001$<br>AIC = 1839.9 | $\Delta R^2 = .009$<br>$F(4) = 9.67, p < .001$<br>AIC = 1836.9 | AIC: Multivariable<br>BIC: PCA |
| **Language** | .280 | 1727.7 | p-tau181<sup>*</sup><br>$\Delta R^2 = .003, p = .036$<br>AIC = 1725.2 | $\Delta R^2 = .003, p = .019$<br>AIC = 1724.1 | $\Delta R^2 = .005$<br>$F(4) = 2.03, p = .088$<br>AIC = 1727.5 | AIC: PCA<br>BIC: Null |
| **Visuospatial** | .339 | 2863.1 | GFAP<br>$\Delta R^2 = .002, p = .059$<br>AIC = 2861.5 | $\Delta R^2 = .000, p = .472$<br>AIC = 2864.5 | $\Delta R^2 = .003$<br>$F(4) = 1.50, p = .199$<br>AIC = 2865.0 | AIC: Individual (GFAP)<br>BIC: Null |

*Note.* Base model (Null) includes age, sex, education, race/ethnicity, APOE $\epsilon 4$ status, and baseline domain score. $\Delta R^2$ = incremental variance explained over the null model. For individual models, the biomarker with the largest $\Delta R^2$ per domain is reported. For the PCA composite, the first unrotated principal component of all four standardized biomarkers was entered as a single predictor (1 df). For the multivariable model, all four biomarkers were entered simultaneously; the omnibus $F$ -test (4 df) tests their joint contribution. AIC = Akaike Information Criterion; BIC = Bayesian Information Criterion. Lower AIC/BIC indicates better fit. AIC favors predictive accuracy; BIC penalizes model complexity more heavily and favors parsimony. The preferred strategy was selected as the model with the lowest AIC or BIC, respectively. Where BIC selects "Null," the biomarker parameter cost was not justified by the improvement in fit. \* Measured in serum; all other biomarkers measured in plasma. *Abbreviations:* R<sup>2</sup>, coefficient of determination; AIC, Akaike Information Criterion; BIC, Bayesian Information Criterion; PCA, principal components analysis; Aβ42/40, amyloid-beta 42/40 ratio; p-tau181, phosphorylated tau-181; GFAP, glial fibrillary acidic protein; NfL, neurofilament light chain

### 3.6 Cross-Sectional-Prospective Differences and Relative Domain Selectivity

#### 3.6.1 Cross-Sectional Versus Prospective Coefficient Differences

Approximate coefficient-difference z-tests comparing standardized multivariable coefficients between the cross-sectional and prospective analyses identified several differences after FDR correction (Supplementary **Table S5**). For p-tau181, coefficients were stronger prospectively than cross-sectionally for memory and global cognition. For Aβ42/40 and NfL, coefficients were stronger cross-sectionally than prospectively for global cognition, with Aβ42/40 also showing a stronger cross-sectional coefficient for executive function. GFAP and NfL also showed borderline FDR-significant coefficient differences for visuospatial ability; because visuospatial associations were not a central finding and primary biomarker associations in this domain were limited, these differences should be interpreted cautiously.

#### 3.6.2 Tertile-Based Associations and Relative Memory-Versus-Executive Selectivity

Tertile-based ANCOVA results for all four biomarkers are displayed in **Figure 2** and Supplementary **Table S6**. Participants in the highest p-tau181 tertile showed lower covariate-and baseline-adjusted cognitive performance at 6-year follow-up for memory (β = −0.237, 95% CI [−0.314, −0.160], q < .001), global cognition (β = −0.213, q < .001), language (β = −0.097, q = .029), and executive function (β = −0.098, q = .005), with no significant visuospatial association (q = .978). In stacked mixed-effects models formally testing relative memory-versus-executive selectivity, the p-tau181 tertile × domain interaction was significant (χ²[2] = 18.50, q < .001). The High-versus-Low tertile effect was larger for memory than for executive function, and the planned Memory − Executive contrast was also significant (estimate = −0.163, SE = 0.039, q < .001).

**Figure 2.**
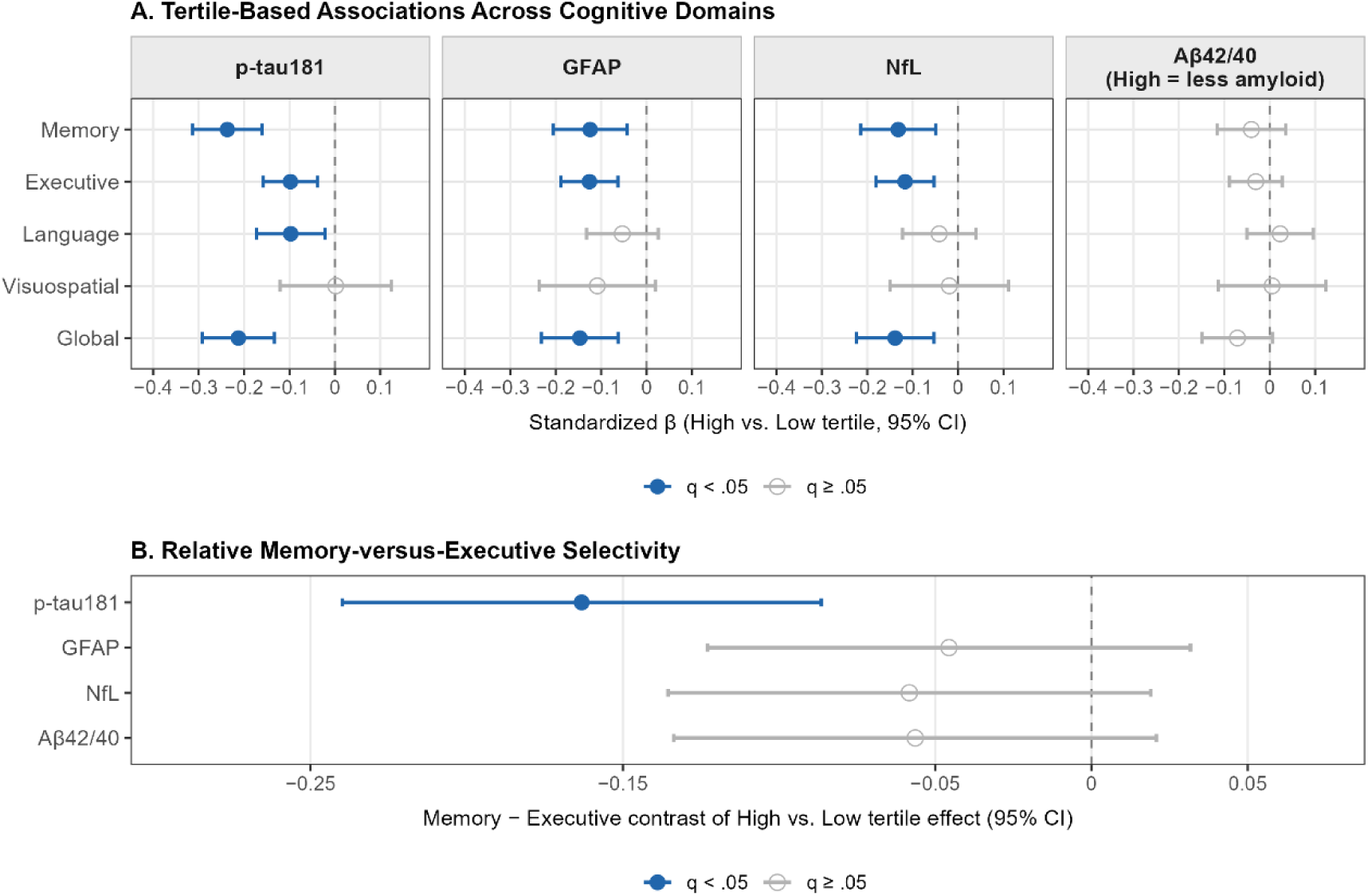
**Panel A.** High-versus-Low tertile associations of each biomarker with baseline-adjusted cognitive performance 6 years later across cognitive domains. Points represent standardized ANCOVA coefficients and error bars 95% confidence intervals; filled points indicate q < .05 after FDR correction. All models adjusted for age, sex, education, race/ethnicity, APOE ε4 status, and the corresponding baseline domain score. For Aβ42/40, higher tertile values indicate lower amyloid burden. **Panel B.** Relative memory-versus-executive selectivity from stacked mixed-effects models. Points represent the Memory − Executive contrast of the High-versus-Low tertile effect; negative values indicate a stronger adverse association for memory than for executive function. *Note.* Tertile groups were defined by biomarker distribution (Low, Middle, High); the High-versus-Low contrast is displayed. Models predict 2022 cognitive performance from 2016 biomarker tertile group membership, adjusted for baseline age, sex, education, race/ethnicity, APOE ε4 carrier status, and the corresponding 2016 domain score. All continuous outcomes were standardized to z-scores. FDR-corrected p-values are reported as q-values. For the memory-versus-executive selectivity analysis (Panel B), the interaction between biomarker tertile and cognitive domain was tested in stacked mixed-effects models with a random intercept for participant; the planned Memory − Executive contrast of the High-versus-Low tertile effect is displayed. **Abbreviations:** ANCOVA, analysis of covariance; FDR, false discovery rate; p-tau181, phosphorylated tau-181; GFAP, glial fibrillary acidic protein; NfL, neurofilament light chain; Aβ42/40, amyloid-beta 42/40 ratio; CI, confidence interval.

Participants in the highest GFAP tertile showed lower memory (β = −0.124, q = .009), executive function (β = −0.126, q < .001), and global cognition (β = −0.147, q = .003), without significant language or visuospatial associations (both q > .24). GFAP showed no evidence of relative memory-versus-executive selectivity (interaction: χ²[2] = 1.66, q = .436; Memory − Executive contrast: estimate = −0.046, SE = 0.039, q = .247). High NfL was similarly associated with memory (β = −0.132, q = .006), executive function (β = −0.117, q = .004), and global cognition (β = −0.139, q = .006), with no significant language or visuospatial associations (both q > .62). NfL also showed no evidence of relative memory-versus-executive selectivity (interaction: χ²[2] = 2.36, q = .436; Memory - Executive contrast: estimate = −0.058, SE = 0.039, q = .203). Aβ42/40 showed no significant prospective tertile associations at any domain (all q > .73), and likewise showed no evidence of relative memory-versus-executive selectivity (interaction: χ²[2] = 2.21, q = .436; Memory − Executive contrast: estimate = −0.056, SE = 0.039, q = .203).

### 3.7 Cross-Sectional and Prospective Composite Analyses

#### 3.7.1 Prospective Composites

Given that p-tau181 and GFAP were the only biomarkers to retain independent prospective associations after mutual adjustment, follow-up exploratory analyses examined whether their additive composite provided a more parsimonious prospective modeling solution. The composite was significantly associated with memory (β = −0.103, q < .001), executive function (β = −0.057, q < .001), language (β = −0.044, q = .010), and global cognition (β = −0.106, q < .001). Model comparison results are presented in **Table 4**. As a single predictor, the composite outperformed the full four-biomarker panel by AIC for memory (ΔAIC = −3.9), language (ΔAIC = −4.9), and global cognition (ΔAIC = −4.3), and by BIC for memory (ΔBIC = −19.1), language (ΔBIC = −20.1), and global cognition (ΔBIC = −19.5). For executive function, the NfL+GFAP composite (ΔBIC = −18.0) and PCA composite (ΔBIC = −17.6) each outperformed the full panel substantially and comparably; the PCA composite was preferred by AIC. The covariate-only null model was preferred by BIC for visuospatial performance at both timeframes.

**Table 4.**
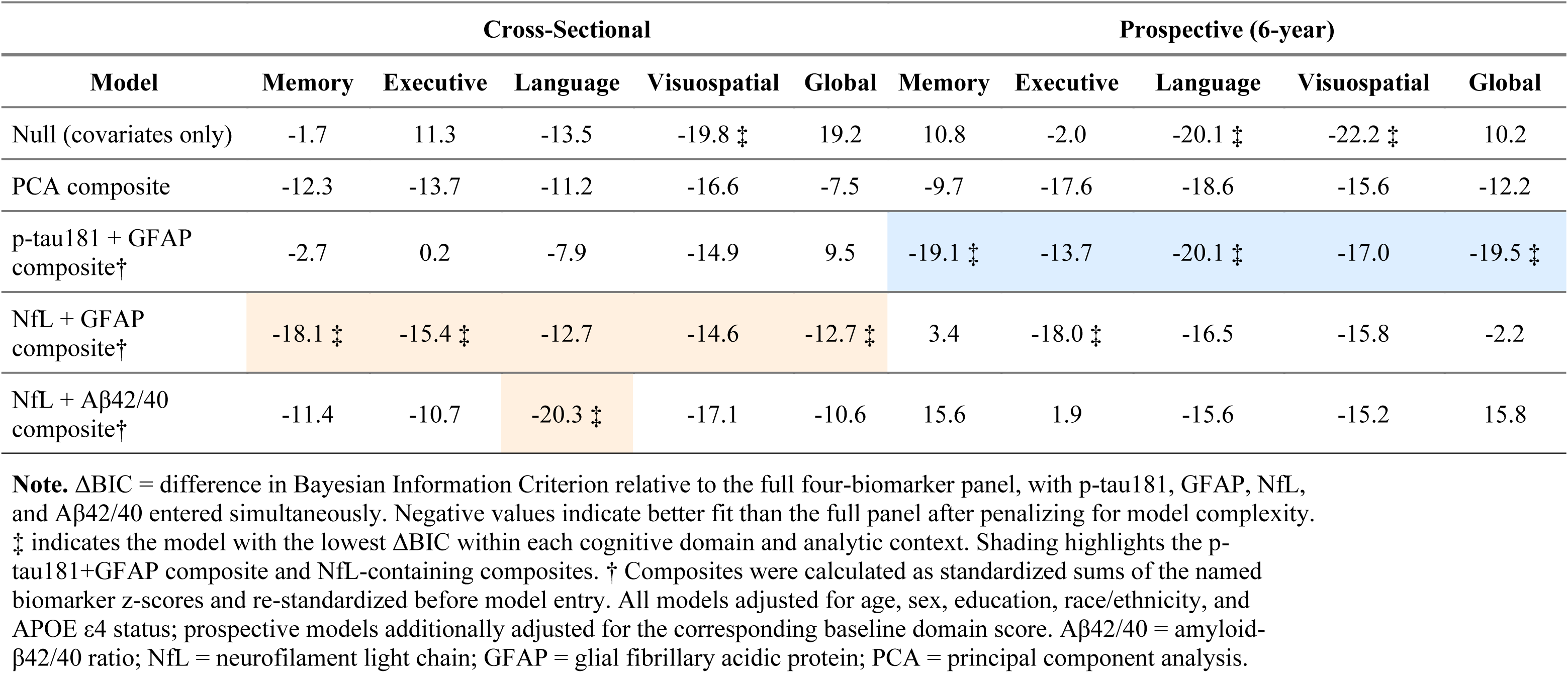
Comparative Fit of Reduced Biomarker Composite Models Relative to the Full Four-Biomarker Panel by Cognitive Domain and Analytic Context.

| Model | Cross-Sectional |  |  |  |  | Prospective (6-year) |  |  |  |  |
| --- | --- | --- | --- | --- | --- | --- | --- | --- | --- | --- |
|  | Memory | Executive | Language | Visuospatial | Global | Memory | Executive | Language | Visuospatial | Global |
| Null (covariates only) | -1.7 | 11.3 | -13.5 | -19.8 ‡ | 19.2 | 10.8 | -2.0 | -20.1 ‡ | -22.2 ‡ | 10.2 |
| PCA composite | -12.3 | -13.7 | -11.2 | -16.6 | -7.5 | -9.7 | -17.6 | -18.6 | -15.6 | -12.2 |
| p-tau181 + GFAP composite† | -2.7 | 0.2 | -7.9 | -14.9 | 9.5 | -19.1 ‡ | -13.7 | -20.1 ‡ | -17.0 | -19.5 ‡ |
| NfL + GFAP composite† | -18.1 ‡ | -15.4 ‡ | -12.7 | -14.6 | -12.7 ‡ | 3.4 | -18.0 ‡ | -16.5 | -15.8 | -2.2 |
| NfL + Aβ42/40 composite† | -11.4 | -10.7 | -20.3 ‡ | -17.1 | -10.6 | 15.6 | 1.9 | -15.6 | -15.2 | 15.8 |

**Note.** ΔBIC = difference in Bayesian Information Criterion relative to the full four-biomarker panel, with p-tau181, GFAP, NfL, and Aβ42/40 entered simultaneously. Negative values indicate better fit than the full panel after penalizing for model complexity. ‡ indicates the model with the lowest ΔBIC within each cognitive domain and analytic context. Shading highlights the p-tau181+GFAP composite and NfL-containing composites. † Composites were calculated as standardized sums of the named biomarker z-scores and re-standardized before model entry. All models adjusted for age, sex, education, race/ethnicity, and APOE ε4 status; prospective models additionally adjusted for the corresponding baseline domain score. Aβ42/40 = amyloid-β42/40 ratio; NfL = neurofilament light chain; GFAP = glial fibrillary acidic protein; PCA = principal component analysis.

#### 3.7.2 Cross-Sectional Composites

Cross-sectional composite analyses identified a different optimal configuration. By BIC (**Table 4**), the NfL+GFAP composite was preferred for memory (ΔBIC = −18.1), executive function (ΔBIC = −15.4), and global cognition (ΔBIC = −12.7), whereas the NfL+Aβ42/40 composite was preferred for language (ΔAIC = −5.1; ΔBIC = −20.3). The p-tau181+GFAP composite, which was preferred prospectively for memory and global cognition, showed only modest improvement over the full panel cross-sectionally for memory (ΔBIC = −2.7) and did not improve fit for global cognition (ΔBIC = +9.5). Conversely, the NfL+GFAP composite, which was preferred cross-sectionally for memory, executive function, and global cognition, did not improve prospective fit for memory (ΔBIC = +3.4), substantially improved fit for executive function (ΔBIC = −18.0), and showed only minimal improvement for global cognition (ΔBIC = −2.2). Overall, the comparatively best-fitting biomarker configuration differed across analytic context, with NfL+GFAP favored for several cross-sectional outcomes and p-tau181+GFAP favored for prospective memory and global cognition. For visuospatial performance, the covariate-only null model was preferred in both analytic contexts.

### 3.8 Sensitivity Analyses

#### 3.8.1 Diagnostic Subgroups

Primary multivariable prospective associations were preserved after excluding participants with baseline dementia (CN + MCI; n = 1,137) and when restricted to cognitively normal participants (CN; n = 931). In both subsamples, p-tau181 remained independently associated with memory (CN+MCI: β = −0.081, q < .001; CN: β = −0.078, q < .001) and global cognition, and GFAP remained associated with executive function, memory, and global cognition. NfL and Aβ42/40 were not independently significant in either subsample. The domain-dependent model preference pattern was also preserved. Full prospective associations for the no-dementia and cognitively normal subsamples are reported in Supplementary **Tables S7** and **S8**, respectively, with corresponding model comparison results in Supplementary **Table S9.**

#### 3.8.2 Race/Ethnicity Moderation

No biomarker × race/ethnicity interaction reached significance after FDR correction (all q > .21; full results in Supplementary **Table S10**). The largest nominal effects were for GFAP × race/ethnicity (F[2] = 4.10, p = .017, q = .294) and p-tau181 × race/ethnicity (F[2] = 3.45, p = .032, q = .294) predicting global cognition, neither of which survived correction.

#### 3.8.3 APOE ε4 Moderation

Three interactions survived FDR correction (Supplementary **Table S11).** NfL × APOE ε4 moderated prospective memory (F[1] = 7.48, p = .006, q = .042): NfL was not associated with memory among non-carriers (β = 0.018, SE = 0.021) but was associated with lower memory among ε4 carriers (β = −0.095, SE = 0.054). GFAP × APOE ε4 moderated prospective executive function (F[1] = 9.71, p = .002, q = .038) and global cognition (F[1] = 7.52, p = .006, q = .042), with larger magnitude associations among ε4 carriers (executive β = −0.120 vs. non-carrier β = −0.029; global β = −0.088 vs. non-carrier β = −0.059). The p-tau181 × APOE ε4 interaction for memory was not significant (q = .651). NfL × APOE ε4 moderation of global cognition was marginally non-significant (q = .059).

## 4 Discussion

This study showed that plasma biomarker–cognition associations in community-dwelling older adults were both domain-differentiated and analytic-context dependent. Cross-sectionally, NfL and GFAP showed the broadest associations with cognition, whereas at six-year follow-up, after accounting for baseline performance and mutual biomarker adjustment, p-tau181 and GFAP carried the most consistent independent signal. P-tau181 also showed relative memory-versus-executive selectivity, and no single biomarker modeling strategy was uniformly optimal across domains. Together, these findings indicate that in population-based cohorts, both the cognitive outcome and biomarker parameterization influence which biomarker–cognition associations are detected and how they are interpreted.

### 4.1 Cross-Sectional and Prospective Dissociation of Biomarker Associations

The dissociation between biomarkers associated with concurrent cognition and those associated with follow-up performance is consistent with evidence that these markers reflect different pathological processes and temporal associations to cognitive decline.^26^ NfL and GFAP, as markers of neuroaxonal injury and astroglial activation, would be expected to track concurrent burden from multiple etiologies, whereas p-tau181 more closely indexes AD-related tau pathology and has been linked to subsequent memory decline in non-demented adults. ^14,50,51^ GFAP’s more consistent associations across analytic contexts is also aligned with prior evidence that it relates to both current cognitive status and future risk for dementia. ^15,52^ Coefficient-difference tests supported this pattern, with p-tau181 showing stronger follow-up than cross-sectional associations for memory and global cognition, whereas NfL and Aβ42/40 showed larger cross-sectional coefficients.

Prior community-based studies have likewise reported domain-differentiated biomarker–cognition associations, although with varying patterns across cohorts. In MYHAT, p-tau181 showed the strongest cross-sectional association with memory, whereas longitudinally Aβ42/40 was most strongly associated with memory change and GFAP with language change ^18^. In HABS-HD, p-tau181 and NfL were each associated cross-sectionally with multiple cognitive domains.^20^ By contrast, shorter follow-up timeframes may be less sensitive to prognostic associations: one study found that baseline plasma p-tau181 was not associated with cognitive or structural brain change over 18 months in cognitively unimpaired older adults ^53^. The present study extends this literature by showing that, even in a heterogeneous community-based sample, p-tau181 retained an independent prospective association with follow-up cognition, with its clearest signal for memory.

### 4.2 Relative Memory-Versus-Executive Selectivity

That p-tau181 was associated with lower memory performance at follow-up is consistent with the known role of medial temporal tau pathology in disrupting episodic memory encoding and consolidation.^16,17^ However, most prior studies have described domain-level patterns by noting which associations reach significance, an approach that does not establish whether a biomarker’s association with one domain differs from its association with another.^23^ In the HABS-HD study, findings showed that p-tau181 was associated with memory, executive function, processing speed, and language but did not formally test whether the associations differed across domains.^20^ By distinguishing true domain-preferential associations from patterns inferred only from statistical significance, such tests can strengthen the clinical interpretation of biomarker profiles and align with recent calls for neuropsychologists to help contextualize blood-based biomarker results.^54^

The absence of relative cognitive selectivity for GFAP and NfL is consistent with their characterization as process markers of astroglial activation and neuroaxonal injury, respectively, neither of which is specific to a single proteinopathy.^2^ GFAP has been associated with executive dysfunction and lower white matter volume in older adults with and without cognitive impairment,^55^ and both markers showed comparable associations across memory and executive function in the present study, as would be expected for markers that index diffuse rather than region-specific pathology.

### 4.3 Modeling Approach and Composite Configuration

Individual biomarker models, composites, and multibiomarker panels have each been used in prior work,^7,12,24,25^ but have rarely been compared systematically within the same cohort against the same cognitive outcomes. In the present study, model performance varied by domain. For follow-up memory, the full multivariable panel was preferred by AIC, whereas BIC favored the more parsimonious individual p-tau181 model; for executive function, the PCA-derived composite was preferred by both criteria. This pattern suggests that the prognostic signal for memory was concentrated primarily in p-tau181, whereas executive function was better captured by variance shared across biomarkers, consistent with the broader and more multifactorial determinants of executive performance, including vascular, inflammatory, and white matter-related processes.^14,55^ Reduced composite analyses supported this interpretation. A p-tau181+GFAP composite outperformed the full panel for follow-up memory, language, and global cognition, indicating that NfL and Aβ42/40 contributed little additional prospective information once p-tau181 and GFAP were considered. Cross-sectionally, by contrast, NfL-containing composites showed the strongest comparative performance for memory, executive function, and global cognition, consistent with the view that concurrent cognition reflects the cumulative burden of neuroaxonal injury from multiple etiologies, a signal that NfL may be particularly sensitive to capture.^6^ Together, these findings suggest that the most informative biomarker representation depends not only on the cognitive domain, but also on whether the analytic aim is concurrent characterization or prospective prediction.

### 4.4 Findings in Cognitively Normal Participants

The persistence of these findings in cognitively unimpaired participants indicates that they were not driven by overt baseline impairment. In these sensitivity analyses, p-tau181 retained its independent association with memory and global cognition, and GFAP retained associations with executive function, memory, and global cognition. These findings are consistent with prior work showing that plasma GFAP carries prognostic information before detectable impairment.^7,15,56^

### 4.5 Moderation by Race/Ethnicity and APOE ε4

Biomarker–cognition associations did not differ by race/ethnicity after FDR correction. Prior work has documented both level differences^9^ and attenuated biomarker–cognition associations in non-Hispanic Black and Hispanic participants relative to non-Hispanic White participants ^20^; however the present sample may not have provided sufficient power to detect such patterns. Three APOE ε4 interactions survived FDR correction: NfL was associated with lower follow-up memory only among ε4 carriers, and GFAP showed stronger associations with executive function and global cognition in ε4 carriers. The p-tau181 × APOE ε4 interaction for memory was not significant. One interpretation is that APOE ε4 may amplify the cognitive influence of more general neurodegenerative and neuroinflammatory processes rather than selectively modifying the tau–cognition association, consistent with prior work,^15^ although differences in outcomes and follow-up duration limit direct comparison.

### 4.6 Limitations

Several limitations should be considered when interpreting these findings. First, the prospective analyses modeled baseline-adjusted follow-up cognition rather than latent change, so they should be interpreted as indexing later cognitive performance conditional on baseline rather than pure cognitive decline. Second, the formal comparisons between cross-sectional and prospective coefficients were based on approximate tests and should therefore be viewed as supportive rather than definitive. Third, some domains, particularly language and visuospatial ability, were represented by fewer measures than memory or executive function, which may have reduced sensitivity for detecting biomarker associations in those domains. Fourth, p-tau181 was measured in serum, whereas GFAP, NfL, and Aβ42/40 were measured in plasma; these matrix and assay differences may have influenced the relative pattern of associations and limit direct comparability across biomarkers. Finally, the absence of PET or CSF measures limits biological validation of the plasma findings, and the composite and moderation analyses should be regarded as exploratory pending replication.

### 4.7 Future Directions

Replication in other population-based cohorts, particularly using models that estimate cognitive change across three or more occasions, would help clarify the extent to which these findings reflect change versus later performance conditional on baseline. Longitudinal biomarker measurement may further determine whether within-person change in p-tau181 and GFAP adds prognostic information beyond baseline levels alone. The observation that a p-tau181+GFAP composite captured much of the informative prospective signal with fewer parameters also raises the possibility that this combination could serve as a more parsimonious prognostic index, which should be tested against incident impairment and dementia outcomes. Finally, direct comparison with p-tau217 and validation in larger cohorts will be important for determining whether the prospective memory signal observed for p-tau181 generalizes to more sensitive tau assays and across subgroups.^3,9,57^

### 4.8 Conclusions

In this population-based cohort of older U.S. adults, plasma biomarkers showed distinct cross-sectional and prospective associations across cognitive domains. NfL and GFAP were most informative for concurrent cognition, whereas p-tau181 and GFAP showed the strongest independent associations with follow-up cognition, and a p-tau181+GFAP composite provided a more parsimonious fit than the full panel for selected outcomes. Only p-tau181 showed formal relative memory-versus-executive selectivity. In sum, these findings support the use of domain-specific cognitive outcomes and biomarker modeling tailored to the analytic context, aligning with recent calls for context-aware biomarker interpretation in heterogeneous community settings.^27^

## Acknowledgments

The authors thank the participants and staff of the Health and Retirement Study-Harmonized Cognitive Assessment Protocol (HRS-HCAP) for their contributions to this research.

## Ethics Statement

The Institutional Review Board of the University of Michigan gave ethical approval for the Health and Retirement Study, from which all human participant data used in this work were obtained. The current study is a secondary analysis of existing Health and Retirement Study data; no additional Institutional Review Board approval was required.

## Consent Statement

All participants provided written informed consent prior to participation in the Health and Retirement Study under a Certificate of Confidentiality from the National Institutes of Health.

## Data Availability Statement

Data used in this study are available through the Health and Retirement Study (https://hrs.isr.umich.edu). Core demographic and cognitive data are available as public-use files. Plasma biomarker, APOE genotype, and HCAP data were accessed through the HRS sensitive data request process. Analytic code is available from the corresponding author upon reasonable request.

